# Wilms tumor mutational subclasses converge to drive *CCND2* overexpression

**DOI:** 10.1101/2023.01.30.23285117

**Authors:** Lin Xu, Kavita Desai, Jiwoong Kim, Qinbo Zhou, Lei Guo, Xue Xiao, Yanfeng Zhang, Li Zhou, Aysen Yuksel, Daniel R. Catchpoole, James F. Amatruda, Kenneth S. Chen

**Affiliations:** Department of Pediatrics, UT Southwestern, Dallas, TX; Quantitative Biomedical Research Center, Department of Population and Data Sciences, Peter O’Donnell Jr. School of Public Health, UT Southwestern, Dallas, TX; Department of Pediatrics, Children’s Hospital of Philadelphia, Philadelphia, PA; Cancer and Blood Disease Institute, Children’s Hospital Los Angeles, Los Angeles, CA; Department of Pediatrics, University of Southern California Keck School of Medicine, Los Angeles, CA; Department of Medicine, University of Southern California Keck School of Medicine, Los Angeles, CA; Australia Biospecimen Research Services, Children’s Cancer Research Unit, Kids Research, The Children’s Hospital at Westmead; Children’s Medical Center Research Institute, UT Southwestern, Dallas, TX

## Abstract

Wilms tumor, the most common kidney cancer in pediatrics, arises from embryonic renal progenitors. Although many patients are cured with multimodal therapy, outcomes remain poor for those with high-risk features. Recent sequencing efforts have provided few biological or clinically actionable insights. Here, we performed DNA and RNA sequencing on 94 Wilms tumors to understand how Wilms tumor mutations transform the transcriptome to arrest differentiation and drive proliferation. We show that most Wilms tumor mutations fall into four classes, each with unique transcriptional signatures: microRNA processing, MYCN activation, chromatin remodeling, and kidney development. In particular, the microRNA processing enzyme *DROSHA* is one of the most commonly mutated genes in Wilms tumor. We show that *DROSHA* mutations impair pri-microRNA cleavage, de-repress microRNA target genes, halt differentiation, and overexpress cyclin D2 (*CCND2*). Several mutational classes converge to drive *CCND2* overexpression, which could render them susceptible to cell-cycle inhibitors.

**Significance:** Through integrated DNA and RNA sequencing analyses, we demonstrate novel genotype-transcriptome associations in Wilms tumors that converge to drive *CCND2* overexpression. Thus, although Wilms tumor mutations are not currently targetable, their effects on the transcriptome may be.

## INTRODUCTION

Wilms tumor is the most common pediatric kidney cancer and one of the most common solid tumors of childhood^1^. Although cure rates for Wilms tumor exceed 90%, multimodal therapy can cause significant long-term therapy, and there are few options for those with high-risk features. In other cancer types, next-generation sequencing has identified targetable mutations. Sequencing studies in Wilms tumor have revealed several recurrent mutations, most commonly *CTNNB1* and *DROSHA*^2-6^. However, these mutations are not therapeutically targetable, and the molecular mechanisms by which many of the most common mutations drive tumor formation remain poorly understood.

Typical “triphasic” Wilms tumors contain three cell types that resemble fetal kidney: blastema, stroma, and epithelia. Just as the metanephric mesenchyme gives rise to stromal and epithelial progenitors during embryonic kidney development, the blastema is thought to give rise to the stromal and epithelial compartments of a Wilms tumor^7^. Because of their histological similarity to fetal kidneys, Wilms tumors are thought to arise from developmental derangements in embryonic renal progenitors. Indeed, many of the genes mutated in Wilms tumor, such as *WT1, SIX1, SIX2*, and *CTNNB1*, are transcription factors for nephron progenitors^5,6^, and the transcriptional program of Wilms tumor resembles that of nephron progenitors^2^. Abnormal methylation at the 11p15 *H19*/*IGF2* locus is also common in Wilms tumor, and it is also seen in precursor lesions known as nephrogenic rests^8^.

We and others previously identified mutations in microRNA processing genes, including *DROSHA, DICER1*, and *DGCR8*, as recurrent driver events in Wilms tumor^2-6^. These mutations impair the production of microRNAs (miRNAs). Loss of miRNAs leads to de-repression of their target genes, such as *LIN28A/B* and *PLAG1*^9,10^. Just as miRNAs were initially discovered to play developmental roles in repressing genes from earlier stages^11,12^, loss of miRNA-mediated gene expression is thought to lock cells into an undifferentiated state. However, the miRNAs and genes responsible for driving the proliferation of renal progenitors remain unknown.

Here, we sought to describe the molecular effects of tumor-driving mutations in 94 Wilms tumors. We performed an integrated multi-omics analysis with targeted capture and whole-genome sequencing, small RNA sequencing, and whole transcriptome sequencing, and we further validated our findings using an independent cohort of previously published sequencing data. Through these analyses, we found that Wilms tumor mutations clustered into four mutational subgroups with unique, direct transcriptional effects. Our findings suggest that a key effect of miRNA loss in Wilms tumor is de-repression of cyclin D2 (*CCND2*), a key regulator of cell cycle progression and well-characterized miRNA target gene. In summary, we found that multiple mutational types converge to drive *CCND2* overexpression in Wilms tumor.

## RESULTS

### Wilms tumor mutations identified by targeted panel, whole-exome, and whole-genome sequencing

To study the effect of mutations in the microRNA processing pathway, we collected Wilms tumor specimens from UT Southwestern and The Children’s Hospital at Westmead in Australia. With Wilms tumor samples from 94 patients, we performed targeted DNA sequencing using a panel of 67 Wilms tumor-related genes and other genes important for miRNA processing. For a subset of tumors, we also carried out whole-exome/whole-genome sequencing, small RNA sequencing, and whole-transcriptome sequencing. We integrated these analyses to understand the molecular and clinical effects of Wilms tumor *DROSHA* mutations.

First, we performed custom targeted deep sequencing of 67 genes in 102 Wilms tumor samples from 94 patients (**Figure 1A**; cases listed in **Supplementary Table S1**; genes listed in **Supplementary Table S2**). In this cohort, we classified mutations into four categories: miRNA processing, MYCN/MAX, chromatin remodeling, and kidney developmental factors. To our knowledge, this type of pathway-driven mutation classification has never been attempted in Wilms tumor. As shown in **Figure 1A**, mutations affecting miRNA processing genes were observed in 19 of 94 primary samples (20%), a proportion consistent with other cohorts. Of these, the most commonly mutated gene was *DROSHA* (mutated in 10 of 94 cases, or 11%). Mutations in miRNA processing factors can overlap with mutations in kidney development genes but are mostly mutually exclusive of other types of mutations.

**Figure 1.**
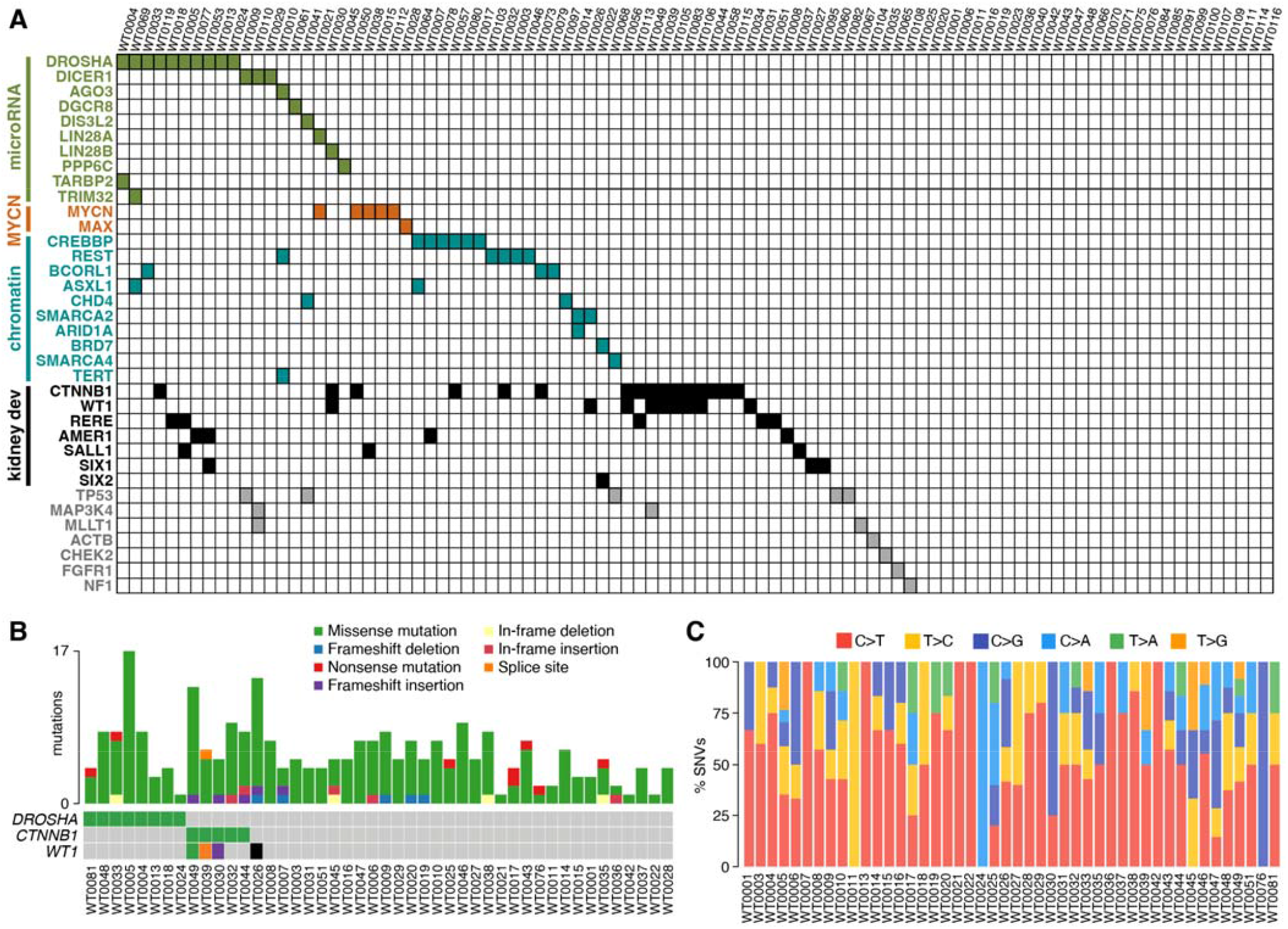
Mutations observed in Wilms tumor. (A) Mutations detected in targeted sequencing panel of Wilms tumor, classified into mutational subclasses. (B) Most commonly mutated genes in tumors analyzed by WES and total number of mutations detected in each tumor. See Supplementary Figure S1 for more details. (C) Most common SNV types in Wilms tumors analyzed by WES.

In addition, for 47 cases where germline tissue and appropriate consent were available, we performed whole-exome sequencing (WES) to identify mutations on a genomic scale. In total, 265 protein-altering mutations were identified, including 246 somatic single-nucleotide variants (SNVs) and 19 somatic small-scale insertion/deletions (indels) (**Supplementary Table S3**). The landscape of recurrent mutated genes among these Wilms tumor patients is shown in **Supplementary Figure S1**. Again, the most frequently mutated gene was *DROSHA* (mutated in 8 of 47 cases, or 17%), and mutations in *DICER1* were observed in an additional 3 cases. Together, mutations in miRNA pathway genes *DROSHA* and *DICER1* were observed in 11 of 47 cases (23%). We also observed recurrent mutations in other previously reported genes, including *CTNNB1* (five patients), *WT1* (four patients), and *AMER1* (three patients) (**Supplementary Figure S1**). Of the 265 mutations identified by WES, 52 were experimentally measured in our custom targeted sequencing panel. All 52 of these mutations were successfully validated by custom targeted deep sequencing (validation rate = 100%, red labels in **Supplementary Table S2**), which confirmed the reliability of our mutation calling.

These tumors in our cohort exhibited a low mutation rate (0.15 non-silent mutations per Mb), which is consistent with previous reports^2,3,5,6^ (**Figure 1B, Supplementary Figure S2A**). Next, we analyzed the mutational signatures based on WES data. We found that the most common somatic SNVs were C>T transitions, followed by T>C transitions (**Figure 1C**). Nearly all the variants were missense SNVs (**Supplementary Figure S2B**). To explore whether the mutation patterns observed here are unique to Wilms tumor, we analyzed TCGA renal cell carcinoma (RCC) WES from with the same computational pipeline to avoid bias. While C>T transitions and missense mutations also predominate in RCC, they are more common in Wilms tumor (**Supplementary Figure S2C**).

Since we failed to identify driver mutations in about one-third of cases, we next generated whole-genome sequencing (WGS) data for 16 tumor-normal pairs to identify somatic SNVs, copy-number alterations (CNAs) and structural variations (SVs) (**Figures 2A-B**; **Supplementary Figure S3; Supplementary Table S4**). As has been previously reported, chromosome 11p, which contains *WT1* and *IGF2*, frequently underwent copy-neutral LOH (WT0035, WT0030, WT0042, and WT0043). Other tumors exhibited loss of chromosome 11 (WT0040), loss of 11p (WT0046), or focal loss of 11p13, the region surrounding *WT1* (WT0029). The most common gain observed was chromosome 1q (n=8 tumors), which is associated with poor outcome^13^, followed by gains of chromosomes 12 and 8 (n=6 and n=5 tumors, respectively). These gains are consistent with previous observations from Wilms tumor chromosomal arrays^13-15^. Consistent with these previous studies, we found that chromosomes 8 and 12 were usually gained together. Chromosome 8 encodes for several genes that may drive tumor formation, including *MYC* and *PLAG1*, and chromosome 12 encodes for both *CCND2* and its binding partner, *CDK4*. WT0028 was the only tumor that exhibited gain(12) without gain(8), but it also harbored a *MAX* mutation that could phenocopy *MYC* gain (**Figure 2B**). These studies suggest that gain of chromosomes 8 and 12 may cooperate in driving Wilms tumor formation. Together, by combining tumor mutational data with copy number changes, we identify driver mutations in 10 of 12 tumors analyzed by WGS.

**Figure 2.**
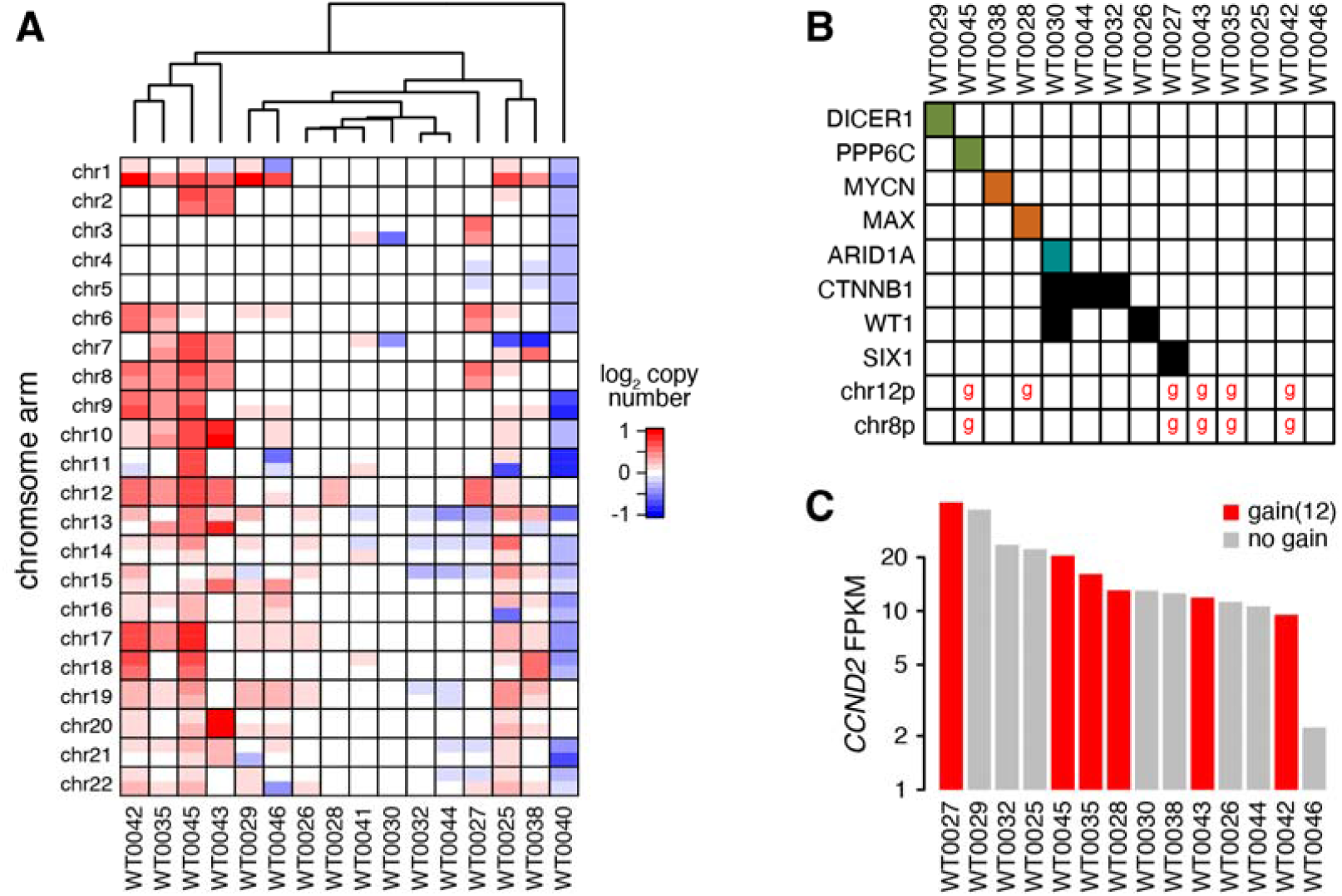
Chromosome 12 gain is associated with high *CCND2* expression. (A) Copy number changes detected by WGS in Wilms tumors. (B) Variants detected by WGS in Wilms tumors. A red ‘g’ represents gain of chromosome 8 or 12. (C) *CCND2* expression in Wilms tumors characterized by WGS.

*CCND2* overexpression has been previously implicated as a Wilms tumor driver^16^. Indeed, previous studies have associated several different genomic features with *CCND2* overexpression, including mutational features associated with our MYCN^17^ and kidney development subgroups^5,18,19^. We confirmed here that tumors in these two subgroups express high levels of *CCND2* (**Figure 2C**). In addition, tumors with an increased copy number of chromosome 12 were among those that expressed high levels of *CCND2*. Lastly, we also found that the two tumors with miRNA pathway mutations also expressed similarly high *CCND2* levels (**Figure 2B-C**).

Although we found putative driver mutations in most tumors, there were many Wilms tumors without a known driver mutation. Since rare cases of translocation-driven Wilms tumors have been reported, we next used fusion detection algorithms to identify inter-chromosomal fusions on tumors that underwent RNA-seq (**Supplementary Table S5**). We detected an *EIF5*-*LIN28B* fusion in WT0071, which causes *LIN28B* expression to be driven by the *EIF5* promoter. This result is consistent with previous reports of translocations involving 6q21, where *LIN28B* resides, in Wilms tumor^13,20-22^.

### Defective microRNA processing in mutant Wilms tumors

We next examined mutations in *DROSHA*, which, along with *CTNNB1*, are the two most commonly mutated genes in Wilms tumor. Although the p.E1147K mutation is the most common *DROSHA* mutation in Wilms tumor, mutations can also arise elsewhere. We compiled previously reported Wilms tumor mutations throughout *DROSHA* from the COSMIC database (**Figure 3A**, which highlights amino acid positions mutated more than once). The DROSHA enzyme processes canonical pri-microRNAs by cleaving their hairpins to produce pre-microRNAs that can be further processed by DICER1. DROSHA has tandem ribonuclease (RNase) III domains, termed RNase IIIa and IIIb, which cleave the 3p and 5p sides of pre-microRNA hairpins, respectively (**Figure 3B**). The *DROSHA* missense mutations seen in Wilms tumor usually affect the metal-binding residues of either RNase III domain. Most mutations affect the RNase IIIb domain, which cleaves on the 5p side, but mutations affecting the RNase IIIa domain are also observed. These mutations affect negatively-charged metal-binding residues (Asp or Glu) in either domain that are critical for ribonuclease function. The only exception is mutations affecting Gln 1187; this charged side-chain protrudes into the same metal-binding pocket and may also be critical for stabilizing a metal ion (**Supplementary Figure S4**).

**Figure 3.**
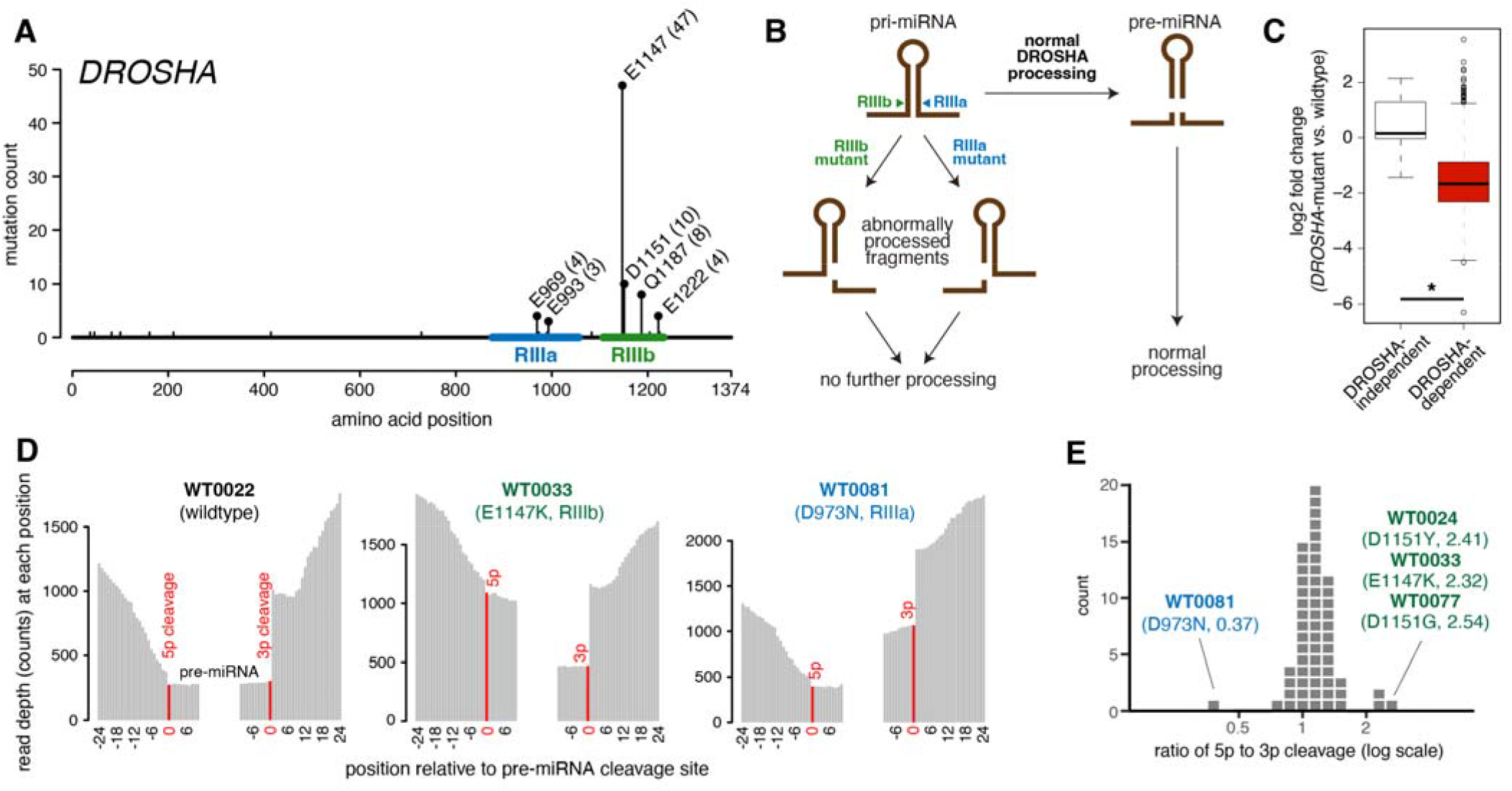
*DROSHA* mutations impair microRNA processing. (A) Wilms tumor mutations in *DROSHA* prior to our study. (B) Patterns of abnormal processing of pri-microRNAs by RIIb- or RIIIa-mutant *DROSHA*. (C) Small RNA sequencing in Wilms tumors with or without mutations in microRNA processing genes, normalized to a spike-in control. (D) Reads mapping to pri-microRNA regions in whole-transcriptome sequencing. (E) Ratio of cleavage at 5p versus 3p sides of pre-microRNAs.

Next, we investigated how *DROSHA* mutations affect miRNA processing using both small RNA sequencing and whole-transcriptome sequencing. First, we performed small RNA sequencing to investigate the effects on miRNA levels. Our previous *in vitro* studies predicted that heterozygous *DROSHA* missense mutations would produce “dominant-negative” global impairment of microRNA processing^3^. These mutations produce abnormally processed fragments that cannot be processed further by DICER1. However, previous studies that measured miRNA levels in Wilms tumor by sequencing did not show such global effects^3,5,6^. To identify global decreases in miRNAs more accurately, we used a spike-in control for absolute normalization. Using spike-in normalization, we observed that tumors with mutations in the miRNA processing pathway exhibited significant miRNA depletion (**Figure 3B-3C, Supplementary Table S6**). Although these mutations are heterozygous, leaving one allele intact, the levels of canonical (*DROSHA*-dependent) miRNAs are depleted further than half. Levels of *DROSHA*-independent miRNAs are unchanged.

Based on our prior work, we expected that a mutation in one domain would still allow processing by the other domain (**Figure 3B**). However, to our knowledge, these partially processed forms have not been observed in human cancer. Thus, we used whole-transcriptome sequencing to examine the effect of *DROSHA* mutations on pri-microRNA processing. Based on our small RNA-seq, we identified miRNAs that were commonly expressed (i.e., expressed in 24 of 32 Wilms tumors) and had well-annotated 5p and 3p ends in miRbase. Correct pri-microRNA processing produces three products: a pre-microRNA, a 5p pri-miRNA arm, and a 3p pri-miRNA arm. However, a mutation in the RIIIb domain, which impairs 5p cleavage, produces only two products; the 5p arm remains connected to the pre-miRNA hairpin. Because pre-microRNAs (60-80 nt) and mature microRNAs (∼20 nt) are too short to be sequenced by standard RNA-seq protocols, tumors with normal miRNA processing have few reads aligning to pre-miRNA regions. Thus, reads aligning to the pre-miRNA region arise from unprocessed or partially processed pri-miRNAs.

We computed the coverage around 5p and 3p pri-miRNA cleavage sites (**Figure 3D**). In tumors with intact pri-miRNA processing, such as WT0022 and WT0035, we observed the expected pattern: an accumulation of reads on the 5p and 3p arms of pre-miRNA hairpins surrounding a flat “valley” representing the pre-miRNA. Our whole-transcriptome sequencing included three tumors with RIIIb mutations (WT0024, WT0033, and WT0077) and one with an RIIIa mutation (WT0081). In tumors with RIIIb mutations, such as WT0024, many more reads span the 5p than the 3p cleavage sites, representing improperly processed pri-miRNAs. Conversely, in WT0081 (RIIIa mutant), reads spanning the 3p cleavage site far outnumbered reads spanning the 5p site. To quantify the efficiency of cleavage on each side of the pri-miRNA hairpin across all tumors, we calculated the ratio of the read depth at the 5p and 3p cleavage sites (**Figure 3E**). Tumors with wild-type *DROSHA* had ratios near 1, whereas the three RIIIb-mutant tumors had ratios greater than 2. Conversely, the only tumor with a ratio less than 0.5 was the RIIIa-mutant tumor.

### Mutational classes correlate with transcriptomic changes

We next analyzed our Wilms tumor whole-transcriptome RNA-seq using gene set enrichment analysis (GSEA)^23^ to study how impaired miRNA processing affects gene expression. We identified the genes predicted to be targeted by commonly expressed microRNAs using TargetScan^24^ predictions, and we used these miRNA target genes as a gene set for GSEA. By comparing tumors with miRNA processing mutations to those without mutations, we verified that these miRNA target genes were enriched in *DROSHA*-mutant Wilms tumors (**Figure 4A**).

**Figure 4.**
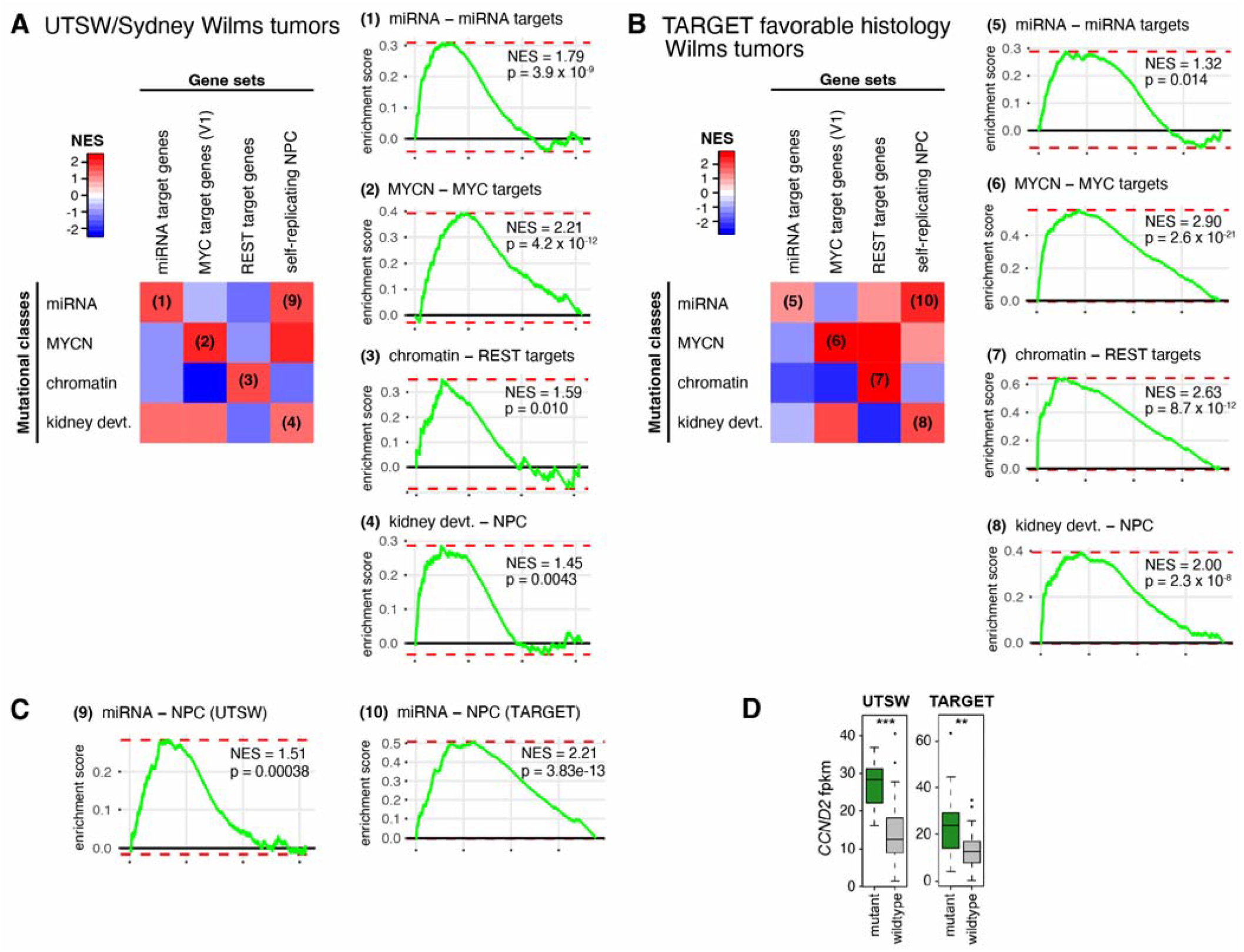
Mutational subclasses of Wilms tumor predict transcriptional changes. (A) GSEA of Wilms tumor samples from our panel. Heatmap (left) shows normalized enrichment score (NES) for each gene set in each of the mutational classes. (B) GSEA of Wilms tumor samples from TARGET study; heatmap shows NES for each gene set in each of the mutational classes. (C) GSEA from both datasets showing enrichment of self-replicating nephron progenitor cell (NPC) signature in miRNA mutational class. (D) *CCND2* expression in our panel and in TARGET Wilms tumors.

Having seen that tumors with mutations in miRNA processing genes correlated with a predictable change in gene expression, we next examined whether other types of mutations also drove transcriptomic changes. Although most tumors contained mutations from only one class, a few contained mutations from more than one class. Thus, we employed a multivariate regression model, treating each mutational class as a categorical variable, to identify how mutational classes correlated with the predicted transcriptional signatures from the Molecular Signatures Database (MSigDB). We also used anaplastic histology as a categorical variable.

As expected, *MYCN* tumors demonstrated enrichment for both sets of MYC target genes as defined by the MSigDB “hallmark” gene set HALLMARK_MYC_TARGETS_V1 (**Figure 4A**). Furthermore, for Wilms tumors driven by mutations in chromatin remodeling genes, the transcriptional repressor *REST* (also known as *NRSF*) was among the most frequently mutated genes in the chromatin remodeling mutational cohort. We found that this class of Wilms tumor exhibited de-repression of *REST* target genes. Out of all “transcription factor targets” gene sets, this was the most enriched signature. Lastly, to assess the effect of the fourth mutational class, kidney developmental genes, we used the self-renewing nephron progenitor cell (NPC) transcriptional signature derived from single-cell RNA-seq of fetal kidneys^25^. We found that tumors with mutations in kidney developmental factors exhibit a gene expression pattern that resembles that of primitive embryonic nephron progenitors, the most undifferentiated population in the developing kidney.

We also examined whether correlations between mutational class and transcriptomic signature were replicated in an independent dataset. We re-analyzed whole-exome sequencing, single-nucleotide polymorphism (SNP) arrays, and RNA-seq from relapsed favorable-histology Wilms tumors from the Therapeutically Applicable Research to Generate Effective Targets (TARGET) project^2^. These analyses were more comprehensive than our dataset, allowing us to identify copy number changes and mutations in genes outside our targeted panel. We re-categorized TARGET tumors into the same mutational classes based on their published mutations and copy number changes (**Supplementary Figure S5**). Based on these mutational classes, we again performed GSEA to examine whether our mutational classification produced the predicted transcriptional changes. Once again, these mutational categories had expected effects on gene expression (**Figure 4B**). Once again, miRNA processing mutations were associated with enrichment of miRNA target genes; *MYCN* alterations with MYC targets; chromatin remodeling mutations with REST targets; and kidney development mutations with nephron progenitor markers. In summary, Wilms tumors have a relatively low mutational burden, and these mutations can lead to widespread effects on the transcriptome.

### Mutated microRNA processing is associated with developmental arrest and *CCND2* overexpression

Having seen that mutational classes correlate with expected transcriptomic signatures, we next explored novel findings that sprung from these signatures. For instance, we noticed that Wilms tumors with mutations affecting microRNA production correlated with a primitive nephron progenitor expression signature in both cohorts (**Figure 4A, 4C**). This finding correlates with the notion that miRNAs can be used during embryonic development to drive differentiation from one state to the next by repressing genes from the prior step. Loss of miRNA processing may contribute to Wilms tumor formation by arresting renal precursors in an undifferentiated state^12^. This shift towards a less differentiated state is consistent with the unique morphological features observed in *DROSHA*-mutant tumors. A previous study associated *DROSHA* mutations with “blastemal-predominant” Wilms tumors^6^, lacking epithelial or stromal components. The NPC signature associated with *DROSHA* mutations may explain this histological observation.

Next, we also examined other hallmark gene sets enriched in these tumors. In our cohort, the hallmark gene set most enriched in tumors with microRNA pathway mutations was HALLMARK_E2F_TARGETS (**Supplementary Figure S6**). These are S-phase genes whose expression is driven by D-type cyclin activity. Thus, we examined changes at the individual gene level, and we found that *CCND2* was expressed at significantly higher levels in *DROSHA*-mutant Wilms tumors, in both UTSW and TARGET cohorts (**Figure 4D**). *CCND2* is well-characterized as a microRNA target gene; its 3′UTR contains several highly conserved binding sites for both miR-16 and let-7 family members^26^. Together, these findings suggest that microRNA loss contributes to Wilms tumor formation through two mechanisms: arresting nephrogenic development and driving proliferation through de-repression of *CCND2*.

## DISCUSSION

In this study, we performed whole-exome, whole-genome, targeted panel sequencing, whole-transcriptome, and small RNA sequencing on a large panel of Wilms tumors. These allow us to find that Wilms tumor mutations affect four classes of genes: miRNA processing, *MYCN* activation, chromatin remodeling, and kidney development. Each of these mutational classes exhibit distinct transcriptional signatures. Such genotype-transcriptome correlations have not previously been described in Wilms tumor. Despite these different transcriptional effects, our analyses suggest that these divergent genomic features converge on the common downstream effect of *CCND2* overexpression to drive Wilms tumor. *CCND2* is known to be highly expressed in ∼80% of Wilms tumors^16,27^, and other studies have associated *CCND2* overexpression with *SIX1/2* mutations^5^, *MYC* activation^17^, Wnt signaling^18^, and *WT1* loss^19^. However, to our knowledge, this is the first report demonstrating that miRNA pathway mutations and gain of chromosome 12 are associated with *CCND2* upregulation in Wilms tumor. Wilms tumor mutations must lock embryonic renal progenitors in an undifferentiated, yet proliferative state, of which *CCND2* may be a key feature.

In particular, little is known about how miRNA pathway mutations drive Wilms tumor; our study suggests that these mutations have two effects: to drive proliferation and to restrain differentiation of renal progenitors. First, we demonstrate that miRNA processing mutations are also associated with *CCND2* overexpression, which drives proliferation. Second, we demonstrate that miRNA processing mutations restrain differentiation through de-repression of NPC marker genes. Because miRNAs commonly repress pluripotency genes during embryonic development^12^, loss of miRNAs are associated with a self-renewing NPC transcriptome.

D-type cyclins control the activity of cyclin-dependent kinases 4 and 6 (CDK4/6), which in turn regulate progression through the G1/S cell cycle checkpoint^28^. Cyclins D1, D2, and D3 can be overexpressed in cancer through amplification, translocation, or loss of the 3′UTR, where microRNAs bind. *CCND2* can be repressed by miR-16, let-7, and other tumor-suppressor microRNAs^26^. Cancers in which D-type cyclins are overexpressed in these three ways are susceptible to CDK4/6 inhibitors *in vitro*^29^. It is thought that *CCND1/2/3* 3′UTR loss drives overexpression by disconnecting cyclins from miRNA regulation. Our study suggests that another way to disconnect D-type cyclins from miRNA regulation is through *DROSHA* mutation. Whether *CCND2* upregulation occurs directly through the loss of miRNA regulation of *CCND2* itself or indirectly through de-repression of factors that regulate *CCND2* transcription remains unknown. Further studies are needed to determine whether Wilms tumors are susceptible to CDK4/6 inhibition.

Compared to previous multi-platform sequencing reports in Wilms tumor, our approach provides certain advantages. First, prior sequencing efforts have not demonstrated an overall decrease in miRNA expression in Wilms tumors with miRNA processing mutations^5,6^. Here, we used a spike-in control to normalize miRNA read counts, which allowed us to detect global changes in miRNA abundance. Similarly, previous efforts have not been able to examine the effects of different types of *DROSHA* mutations on pri-miRNA cleavage activity using RNA-seq. The polyadenylation enrichment used in prior Wilms tumor RNA-seq studies can discard the abnormal products of pri-miRNA cleavage that are not polyadenylated. To our knowledge, this is the first report to document changes in pri-miRNA cleavage in human tumors. Despite the comprehensive nature of our analysis, about many Wilms tumors still lack an obvious driver mutation. Our analysis was not designed to identify changes in methylation, such as imprinting changes at the 11p15 locus, that are not thought to be sufficient for tumorigenesis^30^. Nevertheless, our integrated sequencing analysis provides a high-resolution description of the copy number changes and mutational signatures in Wilms tumor. Future studies may identify driver mutations in noncoding regions or genes that are mutated less commonly that beyond our ability to detect here.

## METHODS

### Wilms tumors from UT Southwestern and The Children’s Hospital at Westmead

Wilms tumor samples were obtained from biospecimen repositories at UT Southwestern and The Children’s Hospital at Westmead. Samples in these repositories were collected at the time of either tumor resection or biopsy, and residual tissue was stored after obtaining informed consent. The study was approved by the Institutional Review Board of the University of Texas Southwestern Medical Center. Genomic DNA was prepared from tumor samples using the DNeasy tissue Kit (Qiagen, Germantown, MD). Small and large RNA were prepared from UTSW samples using the miRNeasy tissue kit (Qiagen). DNA and RNA from The Children’s Hospital at Westmead were prepared using the DNeasy and RNeasy tissue kits (Qiagen).

### Targeted gene sequencing

We chose the panel of genes to be sequenced based on previously reported recurrent mutations in Wilms Tumors ^3,31-34^. Custom capture probes were designed to cover all exons and splice junctions. A total of 94 samples passed quality check. Library preparation and sequencing (100-bp paired-end, ≥1500x depth) were performed at BGI America (Cambridge, MA).

The analysis workflow was based on Genome Analysis Toolkit (GATK, v3.8-0)^35,36^ best practices. Sequencing quality was evaluated using NGS QC Toolkit (v2.3.3)^37^, and high-quality reads were mapped to the UCSC hg19 reference genome using Burrows-Wheeler Aligner (BWA, v0.7.15a)^38^. Picard (v2.12.0) (https://broadinstitute.github.io/picard) was used to remove PCR duplicates, and GATK was used to recalibrate base qualities. Calling variants and genotyping were performed using HaplotypeCaller and the variant calls were filtered using the following criteria: QD (Variant Confidence/Quality by Depth) < 2, FS (Phred-scaled p-value using Fisher’s exact test to detect strand bias) > 60, MQ (RMS Mapping Quality) < 40, DP (approximate read depth) < 3, GQ (Genotype Quality) < 7. The variants were annotated using a custom Perl script (https://github.com/jiwoongbio/Annomen) with mouse transcripts, proteins, and variations (RefSeq and dbSNP build 150).

### Small RNA sequencing

Small RNA libraries were prepared from 32 Wilms tumor samples with adequate RNA. For small RNA, libraries were prepared and sequenced at DNALink, Inc. using the NEBNext Small RNA Library Prep Kit for Illumina, and reads were sequenced on the Illumina NextSeq 500 platform. Trimmed reads were mapped to miRbase using miRDeep2 and normalized to the spike-in control. Differential expression analysis was performed using DESeq2.

### Whole transcriptome sequencing and pri-microRNA processing analysis

Fifty-six tumors and three normal kidney samples underwent whole-transcriptome sequencing. For whole transcriptome sequencing, libraries were prepared using the TruSeq Stranded Total RNA Library Prep Kit (Illumina) and sequenced on the Illumina NovaSeq 6000 platform at DNALink, Inc. 100-bp paired-end reads were assessed for quality, and reads were mapped using CASAVA (Illumina). The generated FASTQ files were aligned by Bowtie2^39^ and TopHat2^40^ to the hg19 human assembly. Cufflinks^41,42^ was used to assemble and estimate the relative abundances of transcripts at the gene and transcript levels.

Differential expression analysis was performed using DESeq2. Specifically, a multivariate linear model was built using each mutational subclass as an independent categorical variable, allowing calculation of the contribution of each mutational subclass to expression of each gene. From these results, the Wald statistic of each gene (calculated as log2-fold-change divided by its standard error) was used for GSEA, using the fgsea package^43^. Unless otherwise stated, gene sets were derived from Molecular Signatures Database (MSigDB v7.1)^44^. Fusion genes were identified by STAR-Fusion (https://github.com/STAR-Fusion/STAR-Fusion) and MapSplice^45^, separately. Only fusion transcripts identified by both algorithms were used in this study.

To study processing of pri-microRNAs, we examined whole-transcriptome sequencing reads that aligned to pri-miRNAs of commonly expressed miRNAs (i.e., microRNAs that were detectable in at least 24 of 32 tumors) whose pri-microRNA cleavage sites (i.e., the first and last position seen in pre-microRNAs) were defined in miRbase. To determine the overall efficiency of 5p and 3p cleavage, we calculated the number of aligned reads around generic 5p and 3p cleavage sites. Ratio of 5p to 3p cleavage efficiency was defined as the ratio of the read depth at the 5p cleavage site to the read depth at the 3p cleavage site.

### Whole-Exome Sequencing and Variant Calling

Exome capture was performed at the UT Southwestern McDermott Next-Generation Sequencing Core using SureSelect Human All Exon v4+UTRs (Agilent Technologies). Sequencing was performed with a HiSeq 2000 instrument (Illumina) with 100-bp paired-end reads. Raw reads were mapped to reference genome (hg38) using BWA ^38^. Duplicates were removed using Picard (http://picard.sourceforge.net/). Local realignment and base quality recalibration were performed using default parameters with the GATK pipeline^46^. Matched tumor-normal BAM files were used as inputs to identify somatic SNVs and indels using the GATK pipeline.

### Whole-genome sequencing, somatic copy number alterations and LOH analysis

Genomic DNA from 16 tumor-normal pairs was analyzed by WGS at the UT Southwestern McDermott Next-Generation Sequencing Core. The WGS data were aligned to the reference genome (hg19) using BWA v.0.7 and sorted by SAMtools v.1.977 and then lifted to hg38. PCR duplicates were removed by Picard (http://picard.sourceforge.net/). By taking matched normal samples as background, tumor-specific copy number alterations and LOH were called by the Somatic Copy-number and Heterozygosity ALteration Estimation (SCHALE)^47^ algorithm using default parameters.

### Statistical analysis

Data analyses were conducted in R v.3.3.2 and Python v.2.7.11 or v.3.5.4. FDR-adjusted *p* values were based on the Benjamini–Hochberg method.

### Re-analysis of TARGET Wilms tumors

Previously reported mutations in Wilms tumor were downloaded from the Catalogue of Somatic Mutations In Cancer (COSMIC v88, https://cancer.sanger.ac.uk/cosmic). These data include mutations previously reported in TARGET Wilms tumors. Recurrently mutated positions in *DROSHA* were identified using this combined source. Next, reported mutations from TARGET tumors were integrated with processed, anonymized Wilms tumor TARGET copy number data and clinical annotations from the TARGET data matrix (https://ocg.cancer.gov/programs/target/data-matrix). At any given gene, tumors were designated as having copy number loss or gain when log2 copy number was <0.67 or >1.65, respectively. Copy number changes at these genes were used for mutational classification: gain of *MYCN* (MYCN); loss of *REST* (chromatin remodeling); and loss of *WT1, AMER1*, or *RERE* (kidney development).

Wilms tumor RNA-seq TARGET data was processed from raw fastq files downloaded from dbGaP. Reads were aligned to the genome (hg38) using Hisat2 (v2.1.0) and assembled using StringTie (v.1.3.2d)^48^ based on the GENCODE v26 reference annotation. Differential gene expression analysis was performed using DESeq2 (v1.36.0)^49^. The four mutational classes (miRNA, MYCN, chromatin remodeling, and kidney development) were used as independent covariates in DESeq2 calculations.

### Data Availability

Sequencing data will be uploaded to dbGAP with accession number XXXX.

## Data Availability

All data produced in the present study will be uploaded to dbGAP upon publication.

## ACKNOWLEDGMENTS

This work was supported by funding from the National Cancer Institute (P50CA196516 to J.F.A., K08CA207849 to K.S.C., and R21CA259771 and P30CA142543 to L.X.), Cancer Prevention and Research Institute of Texas (RR180071 to K.S.C., RP180805 to L.X.), and the Rally Foundation (to L.X.). We thank the patients and families who contributed to this study. We wish to acknowledge the Texas Advanced Computing Center for providing HPC, visualization and other resources that have contributed to the research results reported in this paper.

## SUPPLEMENTARY FIGURES AND TABLES

**Supplementary Table S1**. Wilms tumor specimens analyzed in this study.

**Supplementary Table S2**. Genes targeted by sequencing panel.

**Supplementary Table S3**. SNVs and indels identified by whole-exome sequencing in Wilms tumors.

**Supplementary Table S4**. Copy number and LOH changes identified by whole-genome sequencing in Wilms tumors.

**Supplementary Table S5**. Fusions identified by STAR fusion.

**Supplementary Table S6**. Small RNA sequencing reads, normalized to spike-in control.

**Supplementary Figure S1**. Landscape of mutations identified by whole exome sequencing.

**Supplementary Figure S2**. (A) Mutation burden in Wilms tumors. (B) SNV types in Wilms tumor compared to renal cell carcinoma. (C) Variant classes in Wilms tumor compared to renal cell carcinoma.

**Supplementary Figure S3**. Copy number changes in select Wilms tumors.

**Supplementary Figure S4**. Structure of DROSHA RNase IIIb metal-binding pocket.

**Supplementary Figure S5**. TARGET favorable histology Wilms tumors, categorized by mutational class.

**Supplementary Figure S6**. Enrichment for E2F target genes in Wilms tumors with microRNA processing gene mutations in our dataset.

## Notes

Financial support: This work was supported by funding from the National Cancer Institute (P50CA196516 to J.F.A.; K08CA207849 to K.S.C.; and R21CA259771 and P30CA142543 to L.X.), Cancer Prevention and Research Institute of Texas (RR180071 to K.S.C.; RP180805 to L.X.), and the Rally Foundation (to L.X.).

The authors declare no potential conflicts of interest.

### Competing Interest Statement

The authors have declared no competing interest.

### Author Declarations

The IRB of UT Southwestern Medical Center gave ethical approval for this work.

