## Supplemental Figures for "Wilms tumor mutational subclasses converge to drive *CCND2* overexpression"

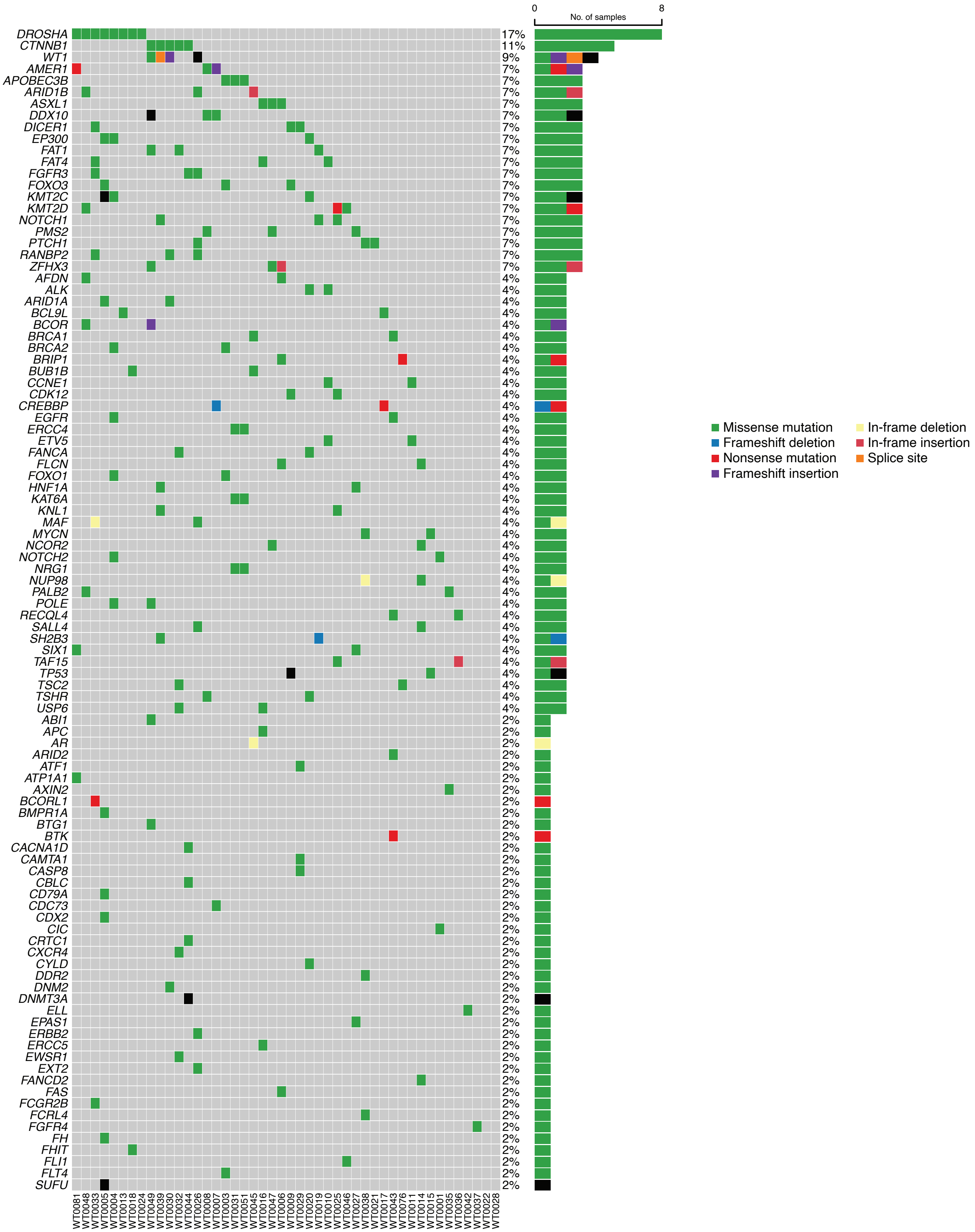


**Supplementary Figure S1.** Landscape of mutations identified by whole exome sequencing.

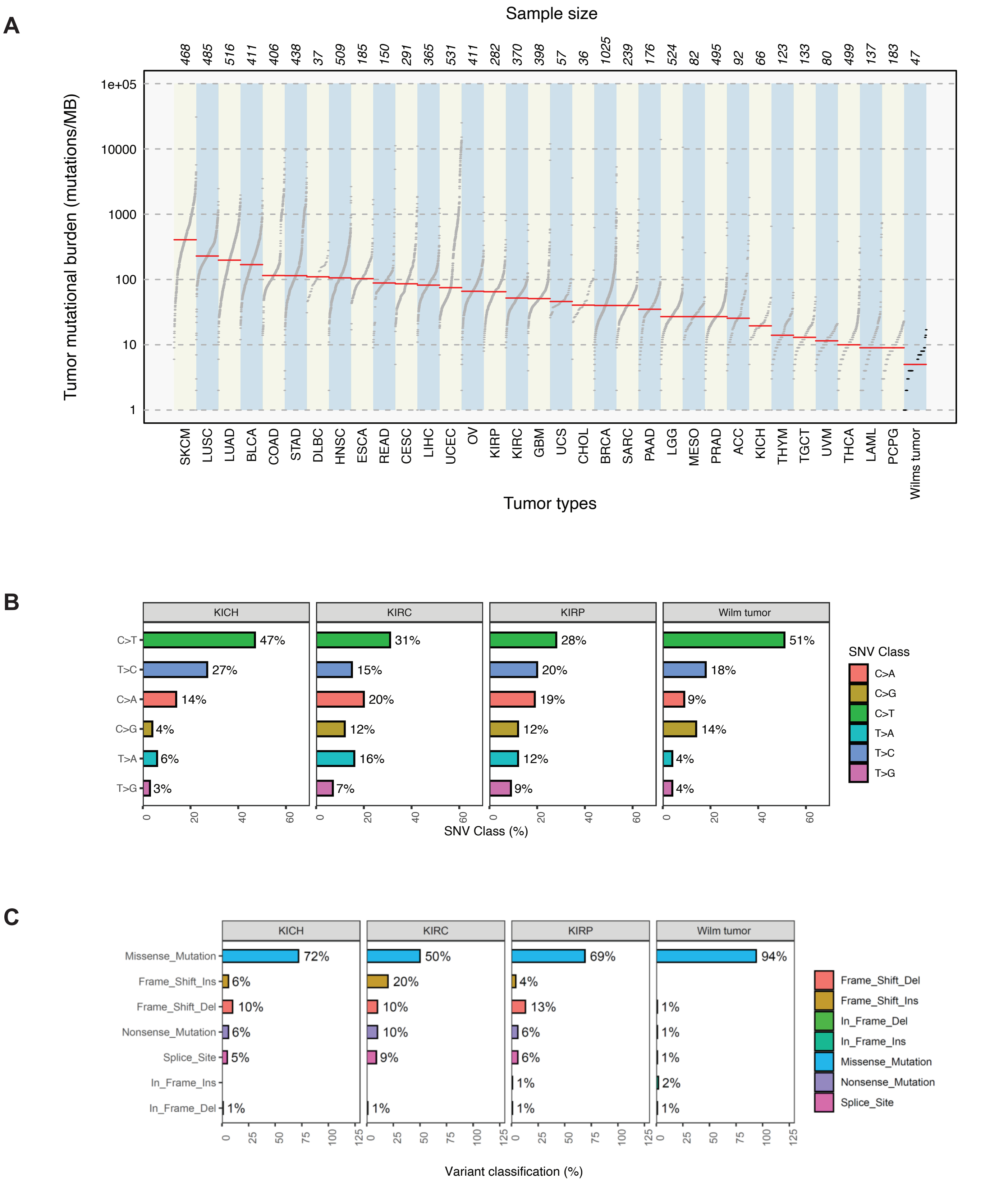


**Supplementary Figure S2.** (A) Mutational burden in Wilms tumors compared to other tumor types in TCGA. (B) SNV types in Wilms tumor compared to subtypes of renal cell carcinoma (RCC; KICH = chromophobe RCC; KIRC = clear cell RCC; KIRP = papillary RCC). (C) Variant classes in Wilms tumor compared to subtypes of renal cell carcinoma.

**A** WT0035

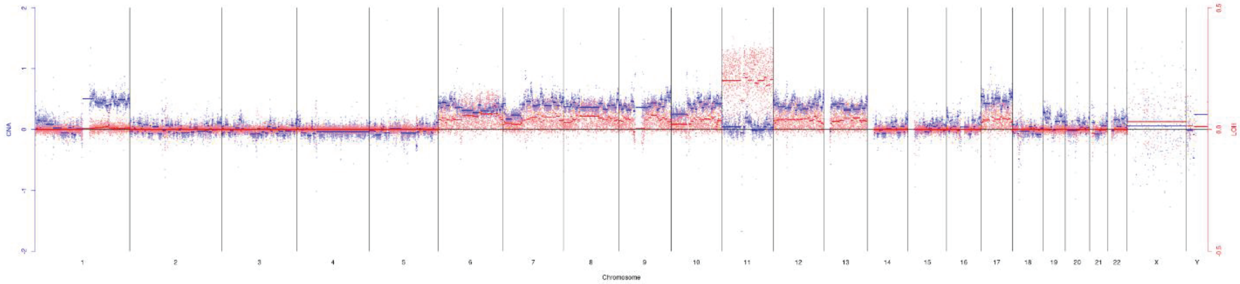


**B** WT0041

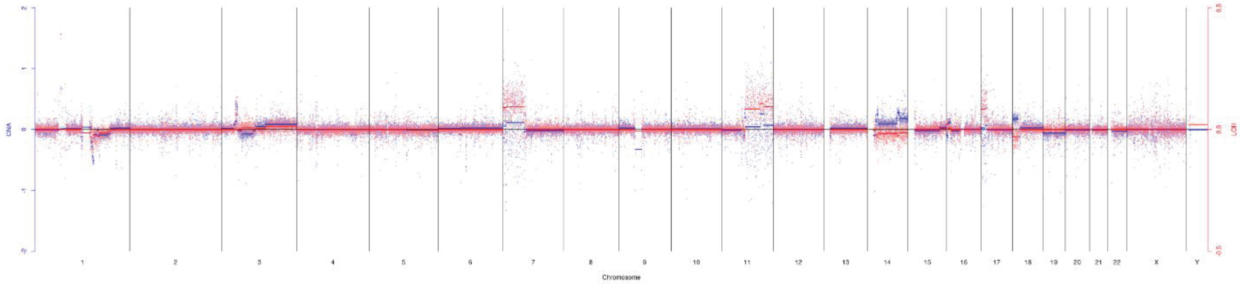


**C** WT0042

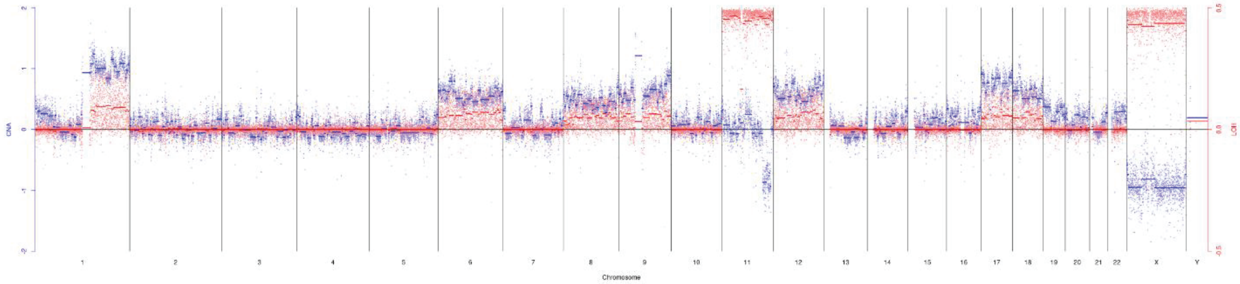


**D** tv

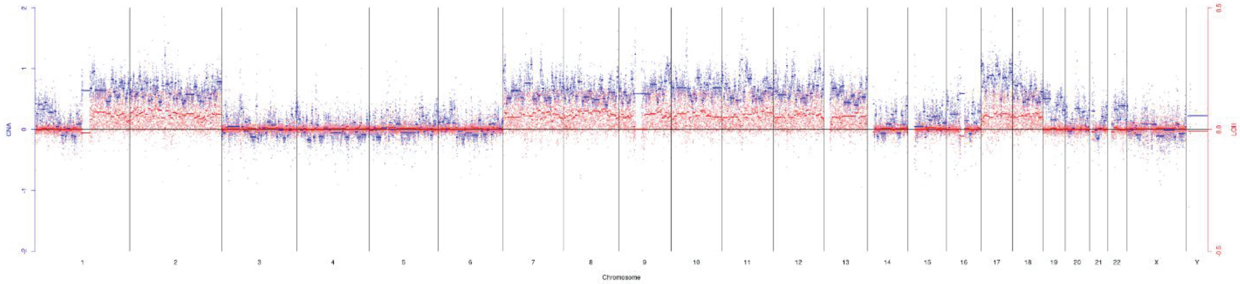


**Supplementary Figure S3.** Copy number changes in select Wilms tumors.

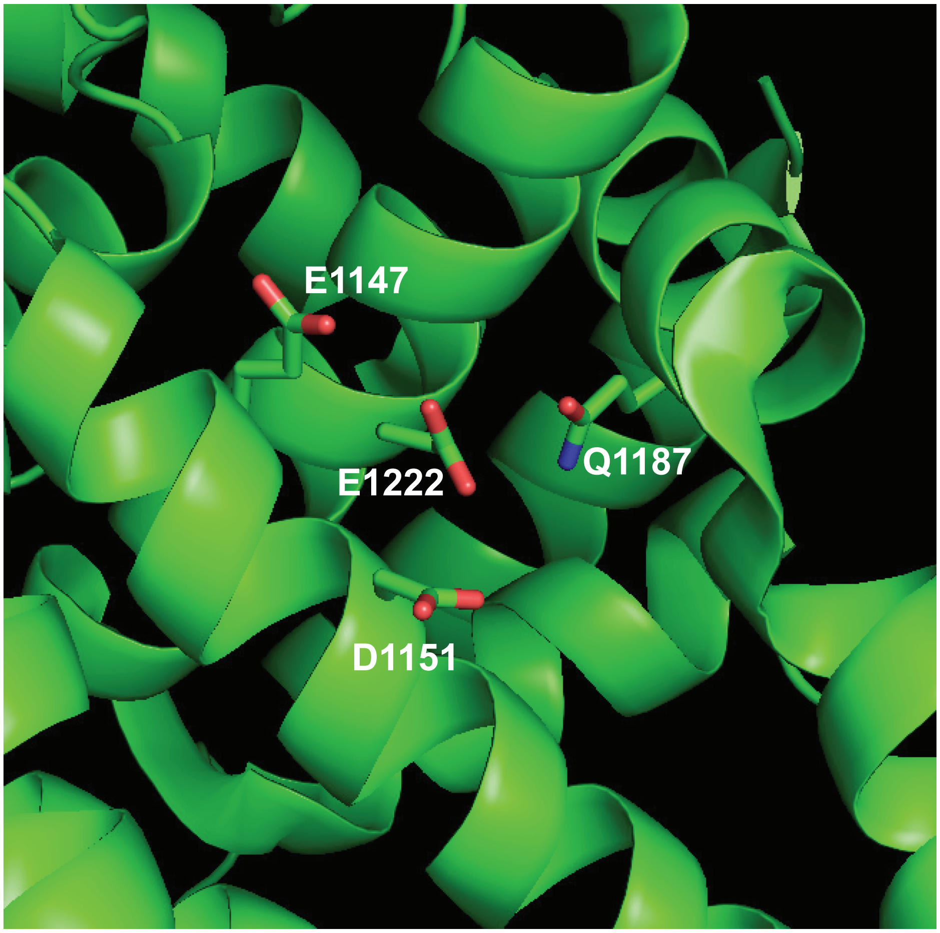


**Supplementary Figure S4.** Structure of DROSHA RNase IIIb metal-binding pocket.

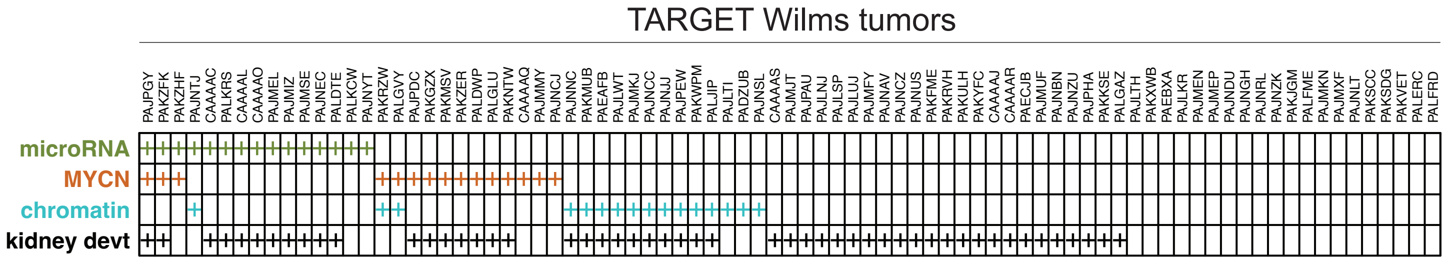


**Supplementary Figure S5.** TARGET favorable histology Wilms tumors, categorized by mutational class.

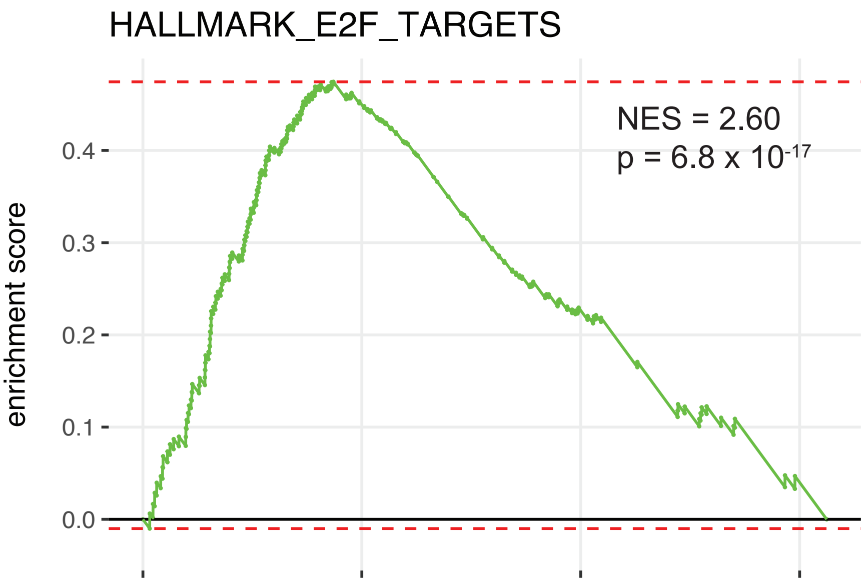


**Supplementary Figure S6.** Enrichment for E2F target genes in Wilms tumors with microRNA processing gene mutations in our dataset.
